# Evaluating signals generated in a large-scale sequence symmetry analyses: macrolides and heart failure; and NSAIDs and pneumonia

**DOI:** 10.1101/2020.03.19.20038596

**Authors:** SJ Sinnott, KJ Lin, S Wang, J Hallas, R Desai, S Schneeweiss, JJ Gagne

**Affiliations:** Division of Pharmacoepidemiology and Pharmacoeconomics, Department of Medicine, Brigham and Women’s Hospital and Harvard Medical School, Boston; Department of Non-Communicable Disease Epidemiology, London School of Hygiene & Tropical Medicine, London; Clinical Pharmacology and Pharmacy, Department of Public Health, University of Southern Denmark, Odense C, Denmark; Department of Clinical Biochemistry and Pharmacology, Odense University Hospital, Odense, Denmark

**Author notes:** **Corresponding author:** Joshua J Gagne, Current. **Authors contributions:** SJS and JJG developed the research questions and wrote the study protocol. SJS carried out all the analyses. SJS and JJG lead the interpretation of the results, with input from KJL, SW, JH, RD and SS. SJS wrote the final manuscript. All authors provided input on the final manuscript and approved the final version.

**Keywords:** Pharmacovigilance, Pharmacoepidemiology, NSAIDs, pneumonia, macrolides, heart failure

## Abstract

**Objective:** Using US claims data and the most up-to-date pharmacoepidemiological study design tools we aimed to investigate two safety signals for (1) macrolides and heart failure; and (2) non-steroidal anti-inflammatory drugs (NSAIDs) and pneumonia generated from a large-scale screening analysis using a self-controlled sequence symmetry design in Danish data.

**Methods:** We used IBM Marketscan data to conduct two new-user, active-comparator cohort studies. In the macrolides example, the exposure was clarithromycin or azithromycin and the comparator was amoxicillin/clavulanate, in patients with sinusitis. In the NSAIDs example, the exposure was oral NSAIDs and the comparator was topical diclofenac, in patients with osteoarthritis. We used Cox proportional hazards regression to estimate hazard ratios (HR) and 95% confidence intervals (CI) to adjust for approximately 50 investigator-specified confounders in a propensity score (PS) matched analysis. In a secondary analysis, we used high-dimensional PS (hd-PS) to adjust for 200 additional proxy confounders.

**Results:** We had 1,012,364 propensity score matched patients exposed to clarithromycin or azithromycin *versus* amoxicillin/clavulanate. With 162 outcomes among clarithromycin or azithromycin exposed patients and 134 among amoxicillin/clavulanate, the HR for overall heart failure was 1.14 (95% CI 0.90 – 1.43). In the NSAIDs example, we included 94,490 patients after propensity score matching. With 794 pneumonia outcomes among oral NSAID patients and 700 among topical diclofenac, we found HR 0.98 (95% CI 0.89 – 1.09). Some upward bias was suspected as larger HRs were observed in the days immediately following exposure for both the macrolides and NSAIDs examples. We found similar results in the hd-PS matched analyses for both examples.

**Conclusion:** Our findings for NSAIDs and pneumonia suggest the original signal may have been due to protopathic or detection bias. Our analyses for macrolides and heart failure with short-term follow-up also suggest bias, although we encourage further research.

## Introduction

The safe and effective use of prescription drugs requires up-to-date and accurate information on adverse events that these drugs can cause. Traditional methods for adverse drug event reporting, such as spontaneous reporting schemes^1^, are limited due to problems with underreporting^2^; lack of denominator data for providing population based incidence estimates; lack of a control group for making causal interpretations; clinical and scientific knowledge required to link drug and unsuspected effects; and sensitivity to widespread publicity.^1^ Thus, alternative methods for identifying adverse drug events in a more systematic manner are warranted.

To this end, we previously used a sequence symmetry analysis (SSA), a self-controlled study design used to detect adverse events associated with drug use, to conduct large-scale hypothesis-free screening in population-level linked data on 450,000,000 drug dispensings and 80,000,000 hospital contacts in Denmark.^3,4^

Our prior screening study found signals for adverse events that are well-known and well understood, such as non-steroidal anti-inflammatory drugs (NSAIDs) and dyspepsia (indicated by use of anti-ulcer drugs). Up to half of the signals we found were a product of reverse causation, such as proton pump inhibitors and dyspepsia.^3^ We also found some associations with unknown mechanisms, such as heart failure occurring after exposure to macrolides, streptogramins, and lincosamides and also pneumonia occurring after exposure to NSAIDs.

The major strengths of the SSA lie in its simplicity in processing and in its robustness to confounders that are stable over time, thereby minimizing the yield of spurious signals caused by this type of confounding.^3^ This, in combination with a growing literature exploring the association between macrolide antibiotics and cardiovascular events^5-9^ indicated that the signal for macrolides, streptogramins and lincosamides and heart failure should be investigated further. In contrast, prior literature examining the relationship between NSAIDs and pulmonary complications in pneumonia has suggested protopathic bias as a possible explanation for the association.^10-12^ Further impetus for focusing on these signals stemmed from macrolide antibiotics and NSAIDs being frequently used drugs across the entire population, thus even a small increase in the risk of heart failure or pneumonia could pose an important public health burden.

We used a new-user, active-comparator design with propensity score-matching within a large administrative claims database, which was different from the database used to conduct the original SSA, to examine whether the associations between (1) macrolides and heart failure; and (2) NSAIDs and pneumonia might represent novel adverse drug event associations.

## Methods

### Data

We used the IBM MarketScan database, which captures longitudinal administrative insurance claims data for a large number of active employees, dependents, retirees and Medicare Supplemental enrollees in the US. The database includes information on plan enrollment, demographics, and integrated records for inpatient events, outpatient events, and pharmacy dispensings.

### Exposure and cohort entry

#### Macrolides and heart failure substudy

The main exposures in the macrolide-heart failure substudy were clarithromycin or azithromycin. The comparator group was amoxicillin/clavulanate. To further minimize confounding by indication, we required all initiators to have a diagnosis code for sinusitis (International Classification of Diseases, Ninth Revision (ICD-9) 461.x, any position in either inpatient or outpatient claims) within 3 days prior to and including the initiation date for the for exposure/comparator drug groups. The rationale for restricting to a sinusitis population was that it would remove individuals treated with macrolides for other indications that may be associated with the heart failure outcome, for example pneumonia.

Patients in the macrolide-heart failure substudy could enter the cohort between 1^st^ Jan 2010 and 30^th^ June 2015. We used this date range to capture an appropriate number of outcomes, thereby facilitating statistical analysis, without having too large a cohort.

#### NSAIDs and pneumonia substudy

For the NSAIDs-pneumonia substudy, the exposure was oral NSAIDs and the comparator was the topical NSAID diclofenac [**APPENDIX 1]**. To further minimize confounding by indication, we required all initiators to have a diagnosis code for osteoarthritis (ICD-9 715.xx^13^, in any position in either inpatient or outpatient claims) in the three-month period prior to and including the initiation date for the exposure/comparator drug groups. We excluded Solaraze™, a topical diclofenac product indicated for actinic keratosis.

Topical diclofenac was approved in the US in 2007. Thus, we allowed cohort entry between 1^st^ Jan 2007 and 30^th^ June 2015 to capture as many topical diclofenac users as possible.

In both examples, the cohort entry date i.e., the index date, was the date on which a patient initiated the study drug or comparator.

### Exclusion criteria

#### Macrolides and heart failure substudy

In the macrolides-heart failure substudy, patients aged <40 years were excluded because younger patients have a low risk of heart failure. We also excluded patients with chronic obstructive pulmonary disease (COPD) or pneumonia measured over all available data prior to cohort entry, because these diagnoses are often differentially associated with users of macrolides vs. other antibiotics in patients with heart failure, which can cause residual confounding.

#### NSAIDs and pneumonia substudy

In the NSAIDs-pneumonia substudy, we excluded patients with contraindications for oral NSAIDs (see list in **Figure 2**).

In both substudies, patients were required to have at least 365 days continuous enrollment in both medical and pharmacy plans prior to cohort entry and no use of either the study or comparator drug during this window. Patients who had new use of both the exposure and the comparator drug on the same date were excluded. Additionally, patients were excluded if they had ever experienced the outcome prior to cohort entry, measured using all available data prior to cohort entry.

### Outcomes

#### Macrolides and heart failure substudy

Heart failure was defined as an inpatient claim in the primary position using ICD-9 code 428.xx, 398.91, 402.01, 402.11, 402.91, 404.01, 404.03, 404.11, 404.13, 404.91, 404.93. This coding algorithm was reported to have 33% sensitivity and 99% specificity in Medicare claims data.^14^

#### NSAIDs and Pneumonia substudy

Pneumonia was defined as an inpatient claim in the primary position or an outpatient claim in any position with any of the following ICD-9 codes; 480.9, 481, 482.xx, 483.x, 484.7, 484.8, 485, 486, 487.0, 488.x1, 517.1.This coding algorithm was derived from a set of validated ICD-10 codes that demonstrated 98% sensitivity and 64% specificity in Australian inpatient data.^15^

### Covariates

#### Macrolides and heart failure substudy

In the macrolide-heart failure substudy we used the following covariates: age categorized as <45years, 45-50years, 50-55years, 55-60years, 60-65years and 65+years; gender; cardiovascular and non-cardiovascular comorbidities; drugs used for prevention of cardiovascular disease and drugs associated with the incidence of heart failure.^16^ All comorbidity and drug variables were constructed using all available data prior to and including the index date.^17^ Patient morbidity and health service utilization were summarized by number of unique drugs, number of inpatient admissions, number of outpatient encounters and a combined comorbidity index all in the 365-day period prior to cohort entry^18^ (**See Table 1 for all covariates)**.

**Table 1:**
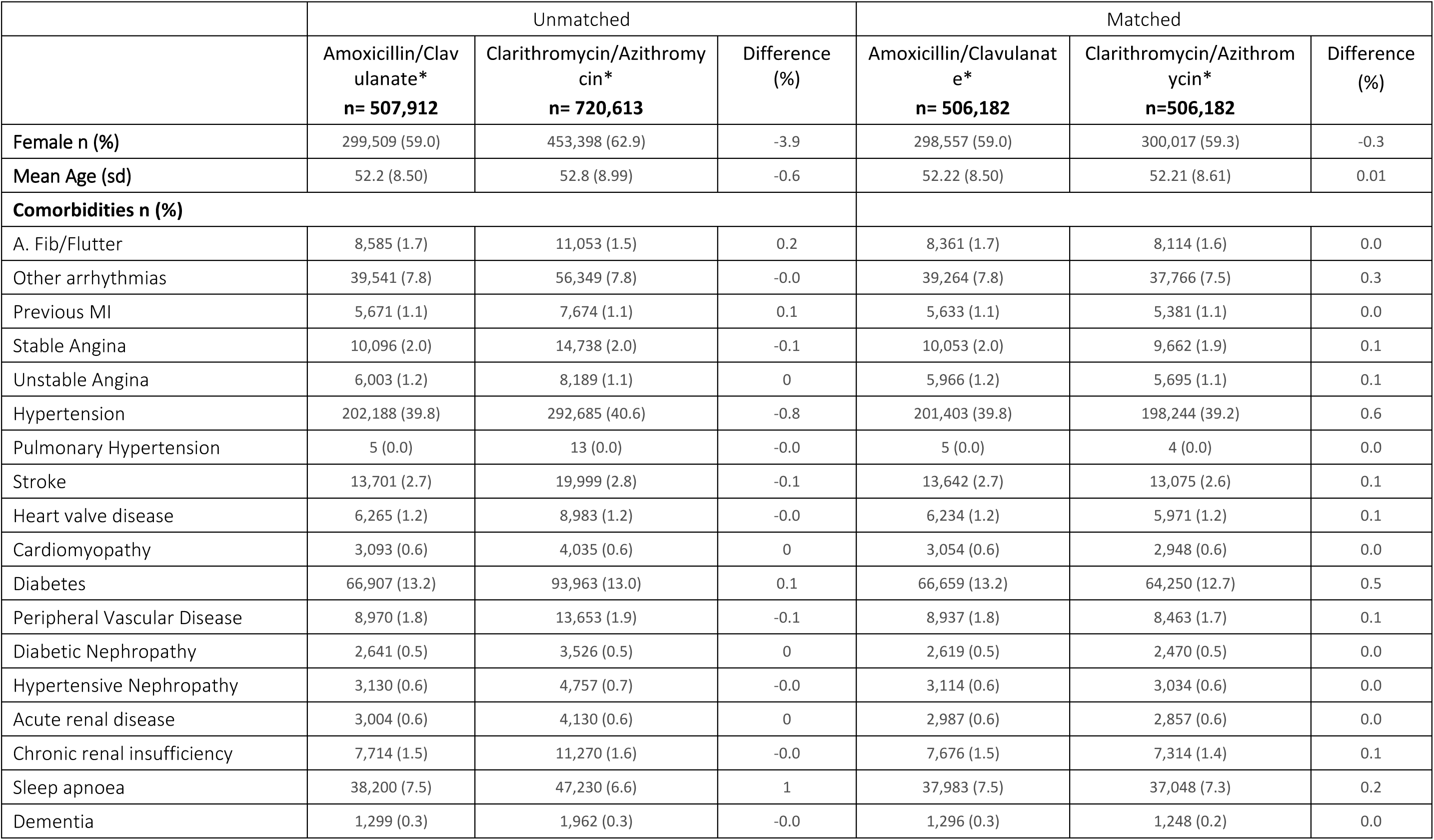
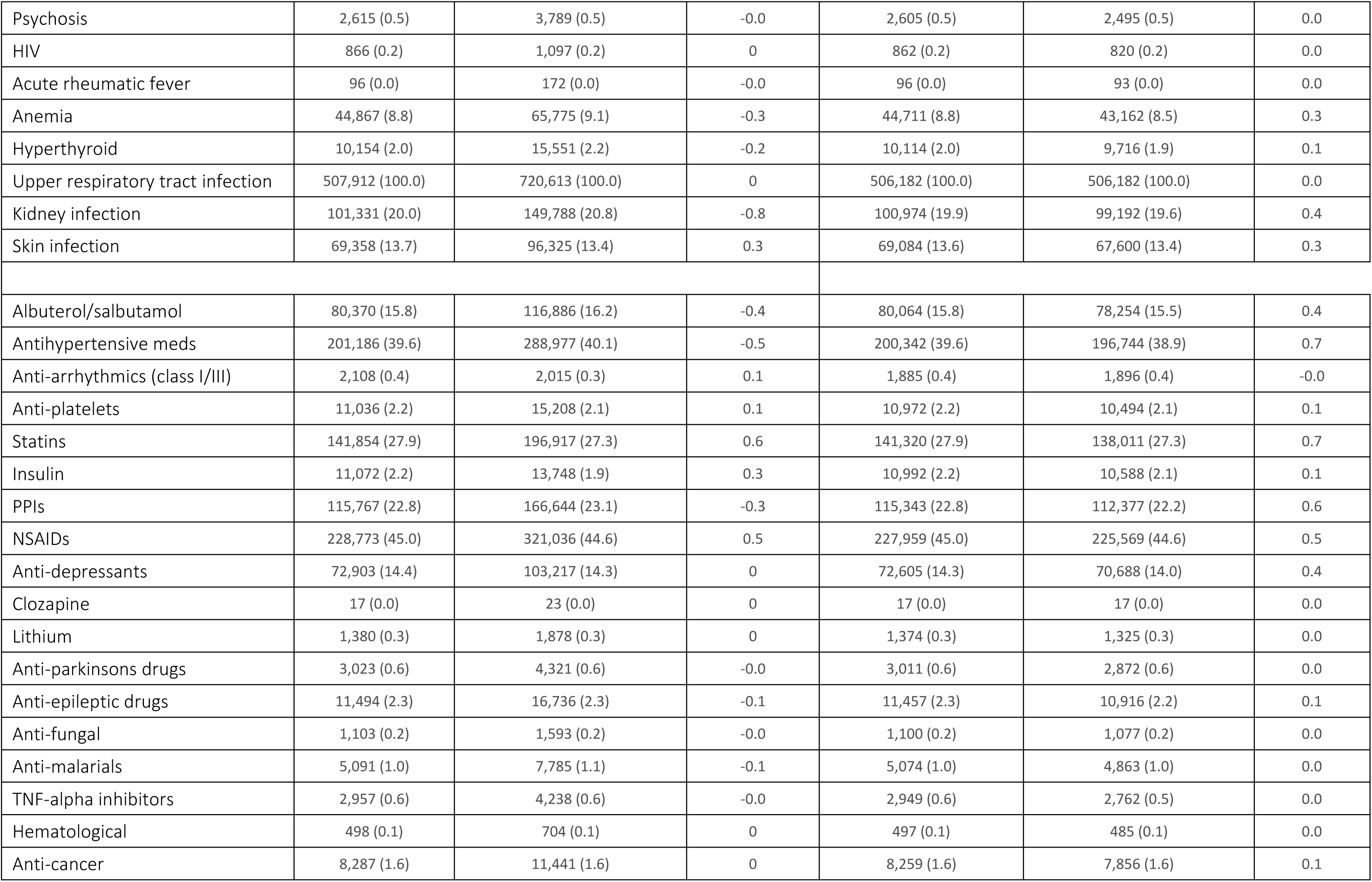
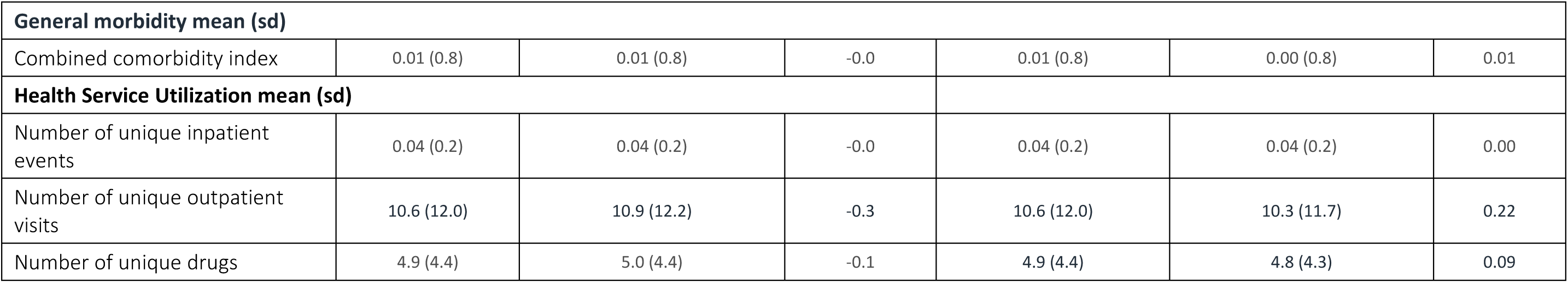
Baseline Characteristics for new initiators of Clarithromycin or Azithromycin (Exposure) and new initiators of Amoxicillin/Clavulanate (comparator)

|  | Unmatched |  |  | Matched |  |  |
| --- | --- | --- | --- | --- | --- | --- |
|  | Amoxicillin/Clavulanate*<br>n= 507,912 | Clarithromycin/Azithromycin*<br>n= 720,613 | Difference (%) | Amoxicillin/Clavulanate*<br>n= 506,182 | Clarithromycin/Azithromycin*<br>n=506,182 | Difference (%) |
| Female n (%) | 299,509 (59.0) | 453,398 (62.9) | -3.9 | 298,557 (59.0) | 300,017 (59.3) | -0.3 |
| Mean Age (sd) | 52.2 (8.50) | 52.8 (8.99) | -0.6 | 52.22 (8.50) | 52.21 (8.61) | 0.01 |
| **Comorbidities n (%)** |  |  |  |  |  |  |
| A. Fib/Flutter | 8,585 (1.7) | 11,053 (1.5) | 0.2 | 8,361 (1.7) | 8,114 (1.6) | 0.0 |
| Other arrhythmias | 39,541 (7.8) | 56,349 (7.8) | -0.0 | 39,264 (7.8) | 37,766 (7.5) | 0.3 |
| Previous MI | 5,671 (1.1) | 7,674 (1.1) | 0.1 | 5,633 (1.1) | 5,381 (1.1) | 0.0 |
| Stable Angina | 10,096 (2.0) | 14,738 (2.0) | -0.1 | 10,053 (2.0) | 9,662 (1.9) | 0.1 |
| Unstable Angina | 6,003 (1.2) | 8,189 (1.1) | 0 | 5,966 (1.2) | 5,695 (1.1) | 0.1 |
| Hypertension | 202,188 (39.8) | 292,685 (40.6) | -0.8 | 201,403 (39.8) | 198,244 (39.2) | 0.6 |
| Pulmonary Hypertension | 5 (0.0) | 13 (0.0) | -0.0 | 5 (0.0) | 4 (0.0) | 0.0 |
| Stroke | 13,701 (2.7) | 19,999 (2.8) | -0.1 | 13,642 (2.7) | 13,075 (2.6) | 0.1 |
| Heart valve disease | 6,265 (1.2) | 8,983 (1.2) | -0.0 | 6,234 (1.2) | 5,971 (1.2) | 0.1 |
| Cardiomyopathy | 3,093 (0.6) | 4,035 (0.6) | 0 | 3,054 (0.6) | 2,948 (0.6) | 0.0 |
| Diabetes | 66,907 (13.2) | 93,963 (13.0) | 0.1 | 66,659 (13.2) | 64,250 (12.7) | 0.5 |
| Peripheral Vascular Disease | 8,970 (1.8) | 13,653 (1.9) | -0.1 | 8,937 (1.8) | 8,463 (1.7) | 0.1 |
| Diabetic Nephropathy | 2,641 (0.5) | 3,526 (0.5) | 0 | 2,619 (0.5) | 2,470 (0.5) | 0.0 |
| Hypertensive Nephropathy | 3,130 (0.6) | 4,757 (0.7) | -0.0 | 3,114 (0.6) | 3,034 (0.6) | 0.0 |
| Acute renal disease | 3,004 (0.6) | 4,130 (0.6) | 0 | 2,987 (0.6) | 2,857 (0.6) | 0.0 |
| Chronic renal insufficiency | 7,714 (1.5) | 11,270 (1.6) | -0.0 | 7,676 (1.5) | 7,314 (1.4) | 0.1 |
| Sleep apnoea | 38,200 (7.5) | 47,230 (6.6) | 1 | 37,983 (7.5) | 37,048 (7.3) | 0.2 |
| Dementia | 1,299 (0.3) | 1,962 (0.3) | -0.0 | 1,296 (0.3) | 1,248 (0.2) | 0.0 |
| Psychosis | 2,615 (0.5) | 3,789 (0.5) | -0.0 | 2,605 (0.5) | 2,495 (0.5) | 0.0 |
| HIV | 866 (0.2) | 1,097 (0.2) | 0 | 862 (0.2) | 820 (0.2) | 0.0 |
| Acute rheumatic fever | 96 (0.0) | 172 (0.0) | -0.0 | 96 (0.0) | 93 (0.0) | 0.0 |
| Anemia | 44,867 (8.8) | 65,775 (9.1) | -0.3 | 44,711 (8.8) | 43,162 (8.5) | 0.3 |
| Hyperthyroid | 10,154 (2.0) | 15,551 (2.2) | -0.2 | 10,114 (2.0) | 9,716 (1.9) | 0.1 |
| Upper respiratory tract infection | 507,912 (100.0) | 720,613 (100.0) | 0 | 506,182 (100.0) | 506,182 (100.0) | 0.0 |
| Kidney infection | 101,331 (20.0) | 149,788 (20.8) | -0.8 | 100,974 (19.9) | 99,192 (19.6) | 0.4 |
| Skin infection | 69,358 (13.7) | 96,325 (13.4) | 0.3 | 69,084 (13.6) | 67,600 (13.4) | 0.3 |
| Albuterol/salbutamol | 80,370 (15.8) | 116,886 (16.2) | -0.4 | 80,064 (15.8) | 78,254 (15.5) | 0.4 |
| Antihypertensive meds | 201,186 (39.6) | 288,977 (40.1) | -0.5 | 200,342 (39.6) | 196,744 (38.9) | 0.7 |
| Anti-arrhythmics (class I/III) | 2,108 (0.4) | 2,015 (0.3) | 0.1 | 1,885 (0.4) | 1,896 (0.4) | -0.0 |
| Anti-platelets | 11,036 (2.2) | 15,208 (2.1) | 0.1 | 10,972 (2.2) | 10,494 (2.1) | 0.1 |
| Statins | 141,854 (27.9) | 196,917 (27.3) | 0.6 | 141,320 (27.9) | 138,011 (27.3) | 0.7 |
| Insulin | 11,072 (2.2) | 13,748 (1.9) | 0.3 | 10,992 (2.2) | 10,588 (2.1) | 0.1 |
| PPIs | 115,767 (22.8) | 166,644 (23.1) | -0.3 | 115,343 (22.8) | 112,377 (22.2) | 0.6 |
| NSAIDs | 228,773 (45.0) | 321,036 (44.6) | 0.5 | 227,959 (45.0) | 225,569 (44.6) | 0.5 |
| Anti-depressants | 72,903 (14.4) | 103,217 (14.3) | 0 | 72,605 (14.3) | 70,688 (14.0) | 0.4 |
| Clozapine | 17 (0.0) | 23 (0.0) | 0 | 17 (0.0) | 17 (0.0) | 0.0 |
| Lithium | 1,380 (0.3) | 1,878 (0.3) | 0 | 1,374 (0.3) | 1,325 (0.3) | 0.0 |
| Anti-parkinsons drugs | 3,023 (0.6) | 4,321 (0.6) | -0.0 | 3,011 (0.6) | 2,872 (0.6) | 0.0 |
| Anti-epileptic drugs | 11,494 (2.3) | 16,736 (2.3) | -0.1 | 11,457 (2.3) | 10,916 (2.2) | 0.1 |
| Anti-fungal | 1,103 (0.2) | 1,593 (0.2) | -0.0 | 1,100 (0.2) | 1,077 (0.2) | 0.0 |
| Anti-malarials | 5,091 (1.0) | 7,785 (1.1) | -0.1 | 5,074 (1.0) | 4,863 (1.0) | 0.0 |
| TNF-alpha inhibitors | 2,957 (0.6) | 4,238 (0.6) | -0.0 | 2,949 (0.6) | 2,762 (0.5) | 0.0 |
| Hematological | 498 (0.1) | 704 (0.1) | 0 | 497 (0.1) | 485 (0.1) | 0.0 |
| Anti-cancer | 8,287 (1.6) | 11,441 (1.6) | 0 | 8,259 (1.6) | 7,856 (1.6) | 0.1 |
| **General morbidity mean (sd)** |  |  |  |  |  |  |
| Combined comorbidity index | 0.01 (0.8) | 0.01 (0.8) | -0.0 | 0.01 (0.8) | 0.00 (0.8) | 0.01 |
| **Health Service Utilization mean (sd)** |  |  |  |  |  |  |
| Number of unique inpatient events | 0.04 (0.2) | 0.04 (0.2) | -0.0 | 0.04 (0.2) | 0.04 (0.2) | 0.00 |
| Number of unique outpatient visits | 10.6 (12.0) | 10.9 (12.2) | -0.3 | 10.6 (12.0) | 10.3 (11.7) | 0.22 |
| Number of unique drugs | 4.9 (4.4) | 5.0 (4.4) | -0.1 | 4.9 (4.4) | 4.8 (4.3) | 0.09 |

#### NSAIDs and pneumonia substudy

In the NSAID-pneumonia substudy we used the following covariates: age categorized as <40years, 40-45 years, 45-50years, 50-55years, 55-60years, 60-65years and 65+years; gender; cardiovascular, respiratory and other comorbidities thought to be risk factors for pneumonia^10,12,19^; drugs used in both cardiovascular, respiratory and other disease states including antibiotic in the past 365 days; and influenza, pneumococcal and herpes zoster vaccines. Patient morbidity and health service utilization were summarized as above, with the inclusion of a frailty index score^20^ (**See Table 1 for all covariates)**.

### Analysis

We used 1:1 propensity score matching to address differences in measured covariates between exposure and comparator groups. We matched the exposure and comparator drug groups by using a nearest-neighbour algorithm and matching caliper of 0.01 on the propensity score. We assessed the performance of the propensity score matching process by evaluating balance in each baseline covariate and overlap in propensity score distributions between treatment groups before and after matching.

We used Cox proportional hazards models to estimate hazard ratios (HR) and 95% confidence intervals (CI) for each outcome in the propensity score-matched cohorts. Follow up started on the index date+1 day for the macrolides-heart failure analysis and started on index date+5 days for the NSAIDS-pneumonia analysis to reduce the potential for protopathic bias (i.e. that NSAIDs were used to treat the earliest manifestations of a yet undiagnosed pneumonia) or detection bias suggested in previous studies.^10,11^ In both analyses, follow-up continued until the earliest of outcome occurrence, end of study period (Sept 30 2015), crossover of exposure group or addition of comparator drug, medical or pharmacy disenrollment, death or 365 days post index date.

### Subgroup analyses

#### NSAIDs and pneumonia substudy

We conducted a subgroup analysis in those who had an ICD-9 code indicating an upper respiratory tract infection **[APPENDIX 2]** in the seven days prior to and including the index date. The rationale was to isolate potential protopathic bias; prior cohort studies have found that young patients with pneumonia are sometimes misdiagnosed as having upper respiratory tract infections and are prescribed NSAIDs. In the days following NSAID exposure, and after respiratory symptoms worsen, the patient is correctly diagnosed as having pneumonia.^10-12^

In a second subgroup analysis we excluded patients with a history of cancer, cirrhosis, COPD, cystic fibrosis, HIV positive status or tracheostomy in the year prior to cohort entry given their increased risk of pneumonia and to help mimic cohorts in prior studies.^10,11^

### Sensitivity analyses

#### Macrolides and heart failure substudy

Brain natriuretic peptide (BNP) test results are commonly used to establish the presence and severity of HF. In a sensitivity analysis, we excluded patients with BNP testing under the assumption that they are not at risk for developing de novo HF.^21^ In a second sensitivity analysis, we redefined the outcome using inpatient codes in any position (83% sensitivity and 86% specificity^14^) to increase numbers of events. Third, we explored clarithromycin only as the exposure drug as opposed to clarithromycin or azithromycin because a clinical trial has examined clarithromycin and it’s association with cardiovascular disease.^6,22^ Fourth, we varied the follow-up period so that follow-up started 7 days after cohort entry and ended 7, 30 and 180 days after cohort entry. We included arrhythmia and myocardial infarction as secondary outcomes to further investigate a potential mechanism underlying an association between the exposure and heart failure.

#### NSAIDs and Pneumonia substudy

We redefined the outcome using only ICD-9 codes (481, 485, 486) that mapped onto the single ICD-10 code (J18) used in the SSA that generated the original signal. Second, we varied follow-up to include the first 5 days of follow-up and, separately, to comprise only the first 5 days of follow-up. We also included a 30- and a 180-day maximum follow-up period. We included community acquired pneumonia as an outcome, defined as any case of pneumonia that was not preceded by a hospitalization of ≥2 days duration in the prior 90 days.^23^

In both substudies we conducted an ‘as treated’ analysis whereby patients were censored at the end of their exposure period. Last, we conducted a high-dimensional propensity score (hdPS) matched analysis for each example.^24^ From drug claims, inpatient claims, outpatient claims, and both inpatient and outpatient procedural claims data we identified and adjusted for 200 variables, additional to pre-specified covariate, based on their prevalence and ability to reduce bias. In the macrcolides-heart failure substudy we excluded ICD-9 code V14 (allergy to penicillin) which may have been highly predictive of macrolide exposure.

### Additional analyses

To investigate the ability of our data and methods to assess known adverse drug events, we explored the association between non-selective NSAIDs and gastrointestinal bleeding, as compared to celecoxib. See **APPENDIX 3** for full details of this analysis.

## Results

### Macrolides and heart failure example

From 185,306,593 individuals available for analysis, 1,228,525 were included in the final cohort for the macrolides-heart failure substudy prior to propensity score matching (**Figure 1)**. In total, 720,613 were exposed to clarithromycin or azithromycin and 507,912 were exposed to amoxicillin/clavulanate. The mean age was 52.5 years (SD 8.8) and the population was 61.3% female. Overall, all covariates were well balanced between the exposure and comparator group. Propensity score matching improved covariate balance even further **(Table 1)**. In the 1:1 propensity score matched cohort, there were 162 heart failure events in 506,182 new initiators of clarithromycin or azithromycin corresponding to a rate of 0.40/1000 person years(py). Median follow-up was 365 days (IQR 235-365). There were 134 heart failure events in the same number of initiators of amoxicillin/clavulanate corresponding to a rate of 0.36/1000py. Median follow-up was 365 days (IQR 182 – 365). In the matched cohort, we did not observe strong evidence to support an increased hazard (HR 1.14, 95% CI 0.90 - 1.43) for heart failure in the clarithromycin or azithromycin group relative to the amoxicillin/clavulanate group.

**Figure 1:**
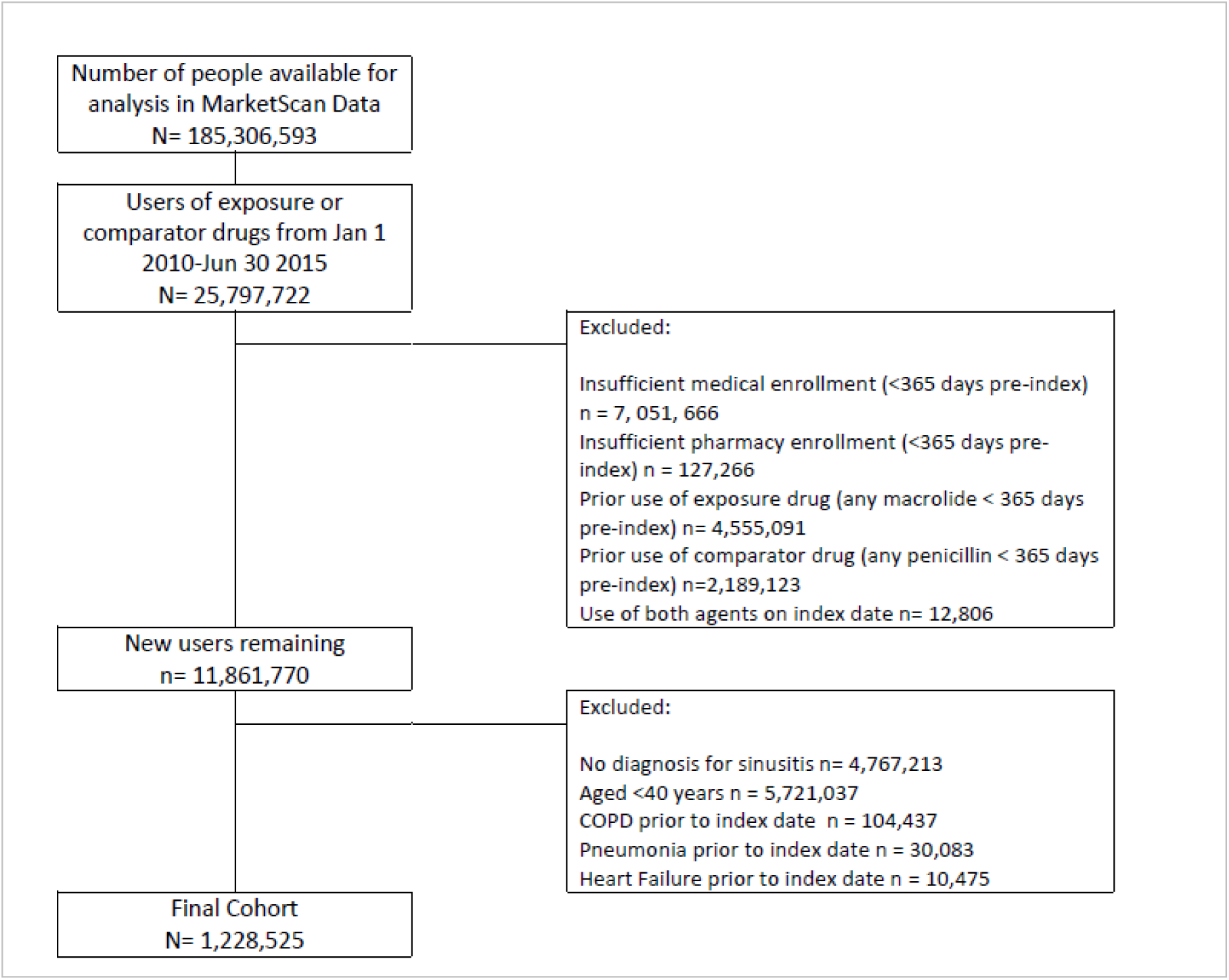
Flowchart describing study cohort inclusion for the heart macrolide-heart failure substudy

### NSAIDs and Pneumonia example

In the NSAIDs-pneumonia substudy 825,923 people were included in the final cohort before propensity score matching (**Figure 2)**. In total, 778,143 were exposed to oral NSAIDs and 47,780 were exposed to topical diclofenac. The mean age was 56.6 years (SD 11.7) and the population was 59.5% female. The prevalence of all comorbidities and drug use was generally higher in the topical diclofenac group. Balance was substantially improved after propensity score matching **(Table 2)**. In the 1:1 propensity score matched cohort, there were 794 pneumonia events in 47,245 new initiators of oral NSAIDs corresponding to a rate of 20.4/1000py. Median follow-up was 365 days (IQR 253 – 365). There were 700 pneumonia events in the same number of new initiators of topical diclofenac corresponding to a rate of 20.8/1000py. Median follow up was 352 days (IQR 145 – 365). In the matched cohort, there was no evidence to support a difference in the occurrence of pneumonia (HR 0.98, 95 CI 0.89 – 1.09) in the exposed group (oral NSAIDs) relative to the topical diclofenac group.

**Figure 2:**
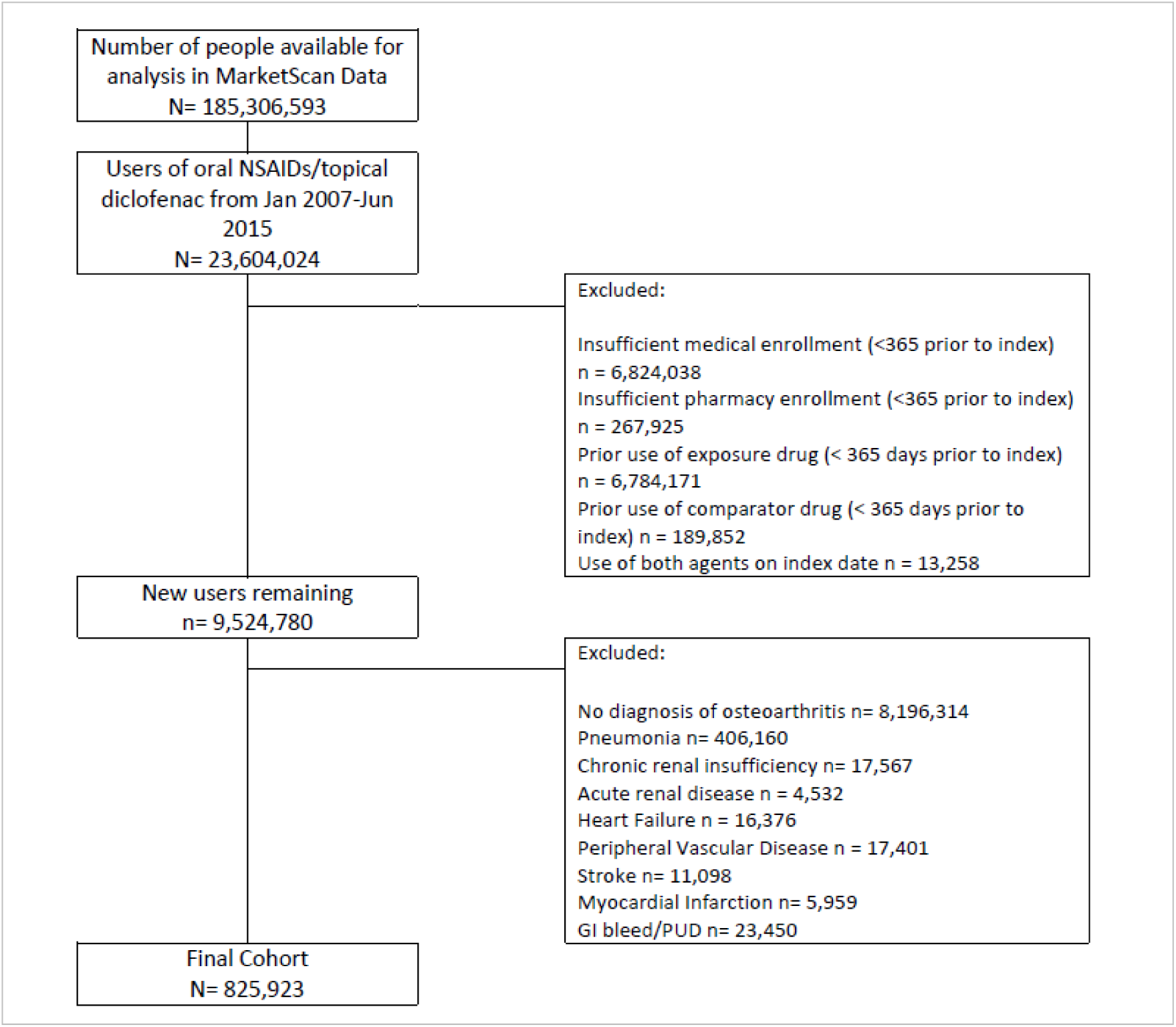
Flowchart describing study cohort inclusion for the NSAIDs–pneumonia substudy

**Table 2:** Baseline Characteristics for new initiators of NSAIDs (Exposure) and new initiators of topical NSAIDs (comparator)

|  | Unmatched |  |  | Matched |  |  |
| --- | --- | --- | --- | --- | --- | --- |
|  | Topical diclofenac<br>n= 47,780 | Oral NSAIDs<br>n= 778,143 | Difference | Topical diclofenac<br>n= 47,245 | Oral NSAIDs<br>n= 47,245 | Difference |
| **Female n (%)** | 34,415 (72.0) | 456,684 (58.7) | 13.3 | 34,044 (72.1) | 34,222 (72.4) | -0.4 |
| **Age mean (sd)** | 60.69 (11.96) | 56.35 (11.68) | 4.34 | 60.71 (11.97) | 60.45 (11.40) | 0.26 |
| **Comorbidities n (%)** |  |  |  |  |  |  |
| Asthma | 6,393 (13.4) | 80,450 (10.3) | 3 | 6,323 (13.4) | 6,238 (13.2) | 0.2 |
| Atrial Fibrillation/Flutter | 2,263 (4.7) | 18,110 (2.3) | 2.4 | 2,245 (4.8) | 2,120 (4.5) | 0.3 |
| Dementia | 435 (0.9) | 4,021 (0.5) | 0.4 | 431 (0.9) | 409 (0.9) | 0 |
| Depression | 6,111 (12.8) | 81,524 (10.5) | 2.3 | 6,029 (12.8) | 6,045 (12.8) | -0.0 |
| Diabetes | 9,423 (19.7) | 136,055 (17.5) | 2.2 | 9,323 (19.7) | 9,260 (19.6) | 0.1 |
| Hypertension | 28,026 (58.7) | 406,857 (52.3) | 6.4 | 27,721 (58.7) | 27,724 (58.7) | -0.0 |
| Parkinsons | 233 (0.5) | 2,230 (0.3) | 0.2 | 230 (0.5) | 219 (0.5) | 0 |
| Stable Angina | 1,661 (3.5) | 20,530 (2.6) | 0.8 | 1,644 (3.5) | 1,644 (3.5) | 0 |
| Unstable Angina | 785 (1.6) | 9,935 (1.3) | 0.4 | 777 (1.6) | 796 (1.7) | -0.0 |
| Upper respiratory tract infection | 404 (0.8) | 8,995 (1.2) | -0.3 | 400 (0.8) | 381 (0.8) | 0 |
| Cancer (exc skin cancer) | 25,027 (52.4) | 326,710 (42.0) | 10.4 | 24,753 (52.4) | 24,830 (52.6) | -0.2 |
| Skin Cancer | 4,335 (9.1) | 48,400 (6.2) | 2.9 | 4,292 (9.1) | 4,242 (9.0) | 0.1 |
| Cirrhosis | 185 (0.4) | 1,614 (0.2) | 0.2 | 183 (0.4) | 154 (0.3) | 0.1 |
| COPD | 2,618 (5.5) | 34,973 (4.5) | 1 | 2,600 (5.5) | 2,661 (5.6) | -0.1 |
| Cystic Fibrosis | 17 (0.0) | 227 (0.0) | 0 | 15 (0.0) | 12 (0.0) | 0 |
| HIV | 52 (0.1) | 959 (0.1) | -0.0 | 50 (0.1) | 52 (0.1) | -0.0 |
| Tracheostomy | 8 (0.0) | 91 (0.0) | 0 | 8 (0.0) | 5 (0.0) | 0 |
| **Drugs n (%)** |  |  |  |  |  |  |
| Inhaled corticosteroids | 17,367 (36.3) | 222,010 (28.5) | 7.8 | 17,190 (36.4) | 17,121 (36.2) | 0.1 |
| Insulin | 1,367 (2.9) | 17,867 (2.3) | 0.6 | 1,357 (2.9) | 1,387 (2.9) | -0.1 |
| Inhaled bronchodilators | 10,050 (21.0) | 135,570 (17.4) | 3.6 | 9,939 (21.0) | 9,872 (20.9) | 0.1 |
| Recent antibiotic | 40,101 (83.9) | 626,887 (80.6) | 3.4 | 39,641 (83.9) | 39,701 (84.0) | -0.1 |
| PPIS | 16,930 (35.4) | 204,316 (26.3) | 9.2 | 16,737 (35.4) | 16,783 (35.5) | -0.1 |
| H2-antagonists | 3,451 (7.2) | 43,829 (5.6) | 1.6 | 3,422 (7.2) | 3,502 (7.4) | -0.2 |
| Statins | 18,905 (39.6) | 263,893 (33.9) | 5.7 | 18,714 (39.6) | 18,728 (39.6) | -0.0 |
| Anti-platelets | 1,516 (3.2) | 15,593 (2.0) | 1.2 | 1,505 (3.2) | 1,494 (3.2) | 0 |
| Anti-diabetes | 5,758 (12.1) | 87,218 (11.2) | 0.8 | 5,705 (12.1) | 5,658 (12.0) | 0.1 |
| Anti-hypertensives | 27,548 (57.7) | 395,412 (50.8) | 6.8 | 27,250 (57.7) | 27,350 (57.9) | -0.2 |
| Benzodiazepines | 15,747 (33.0) | 197,796 (25.4) | 7.5 | 15,557 (32.9) | 15,664 (33.2) | -0.2 |
| Opioids | 17,694 (37.0) | 247,856 (31.9) | 5.2 | 17,504 (37.0) | 17,514 (37.1) | -0.0 |
| **Vaccines n (%)** |  |  |  |  |  |  |
| Influenza | 1,946 (4.1) | 22,033 (2.8) | 1.2 | 1,920 (4.1) | 1,923 (4.1) | -0.0 |
| Pneumococcal | 97 (0.2) | 939 (0.1) | 0.1 | 95 (0.2) | 78 (0.2) | 0 |
| Zoster | 1,484 (3.1) | 11,403 (1.5) | 1.6 | 1,467 (3.1) | 1,401 (3.0) | 0.1 |
| **General Morbidity mean (sd)** |  |  |  |  |  |  |
| Frailty Index | 0.1 (0.04) | 0.1 (0.04) | 0.01 | 0.1 (0.04) | 0.1 (0.04) | 0.00 |
| **Health Service Utilization in year prior to cohort entry mean (sd)** |  |  |  |  |  |  |
| Number of unique inpatient events | 0.1 (0.4) | 0.1 (0.4) | -0.00 | 0.1 (0.4) | 0.1 (0.4) | 0.00 |
| Number of unique outpatient visits | 20.5 (17.5) | 14.9 (14.2) | 5.57 | 20.5 (17.5) | 20.3 (18.3) | 0.19 |
| Number of Unique Drugs | 8.3 (5.8) | 6.5 (5.1) | 1.82 | 8.3 (5.8) | 8.3 (5.8) | 0.00 |

## Subgroup Analyses

### NSAIDs and pneumonia example

In patients with a diagnosis code for an upper respiratory tract infection in the week prior to and including index date we observed HR 1.50 (95% CI 0.59 – 3.82). In a population that was restricted by strong risk factors for pneumonia we found similar results to the main analysis that had adjusted for these risk factors **(Figure 4)**.

## Sensitivity Analyses

### Macrolides and heart failure example

We re-defined the outcome using inpatient claims in any position as opposed to the primary position and found a similar risk of heart failure as the main analysis (HR 1.09, 95% 0.98 – 1.21). We removed patients with prior BNP testing (indicating uncoded/undiagnosed HF) and again found similar risk as the main analysis (HR 1.17. 95% CI 0.93 – 1.48) (**Figure 3)**. When the exposure was defined as clarithromycin only, the confidence limits overlapped with those of the main analysis **(Figure 3)**. We did not find evidence to support an association between clarithromycin or azithromycin and myocardial infarction (HR 1.06, 95% CI 0.93 – 1.20) or arrhythmia (HR 0.96, 95% CI 0.85 – 1.08). We found that the hazard ratio was strongest in the seven day period immediately following exposure (HR 3.49, 95% CI 1.15 – 10.60) and weakest when the first seven days of follow-up were removed (HR 1.07, 95% CI 0.85 – 1.35). An hdPS matched cohort yielded results very similar to the main analysis (**Figure 3)**.

**Figure 3:**
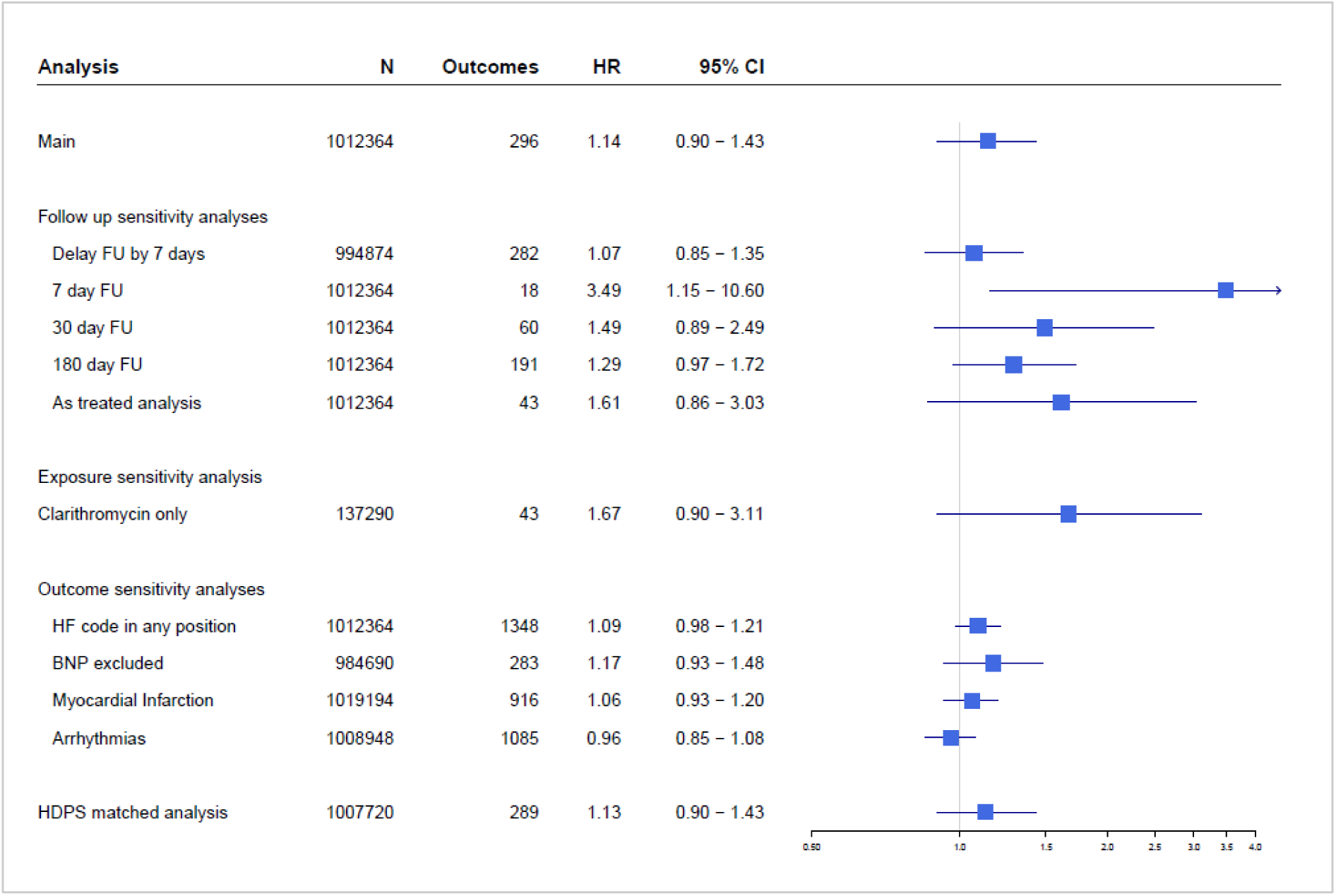
Forest plot demonstrating hazard ratios and 95% CI for the main macrolides - heart failure analysis and all subgroup and sensitivity analyses. Main exposure was clarithromycin or azithromycin in sinusitis, comparator was amoxicillin/clavulanate in sinusitis.

### NSAIDs and pneumonia example

An analysis of only the first 5 days of follow-up gave an increased hazard for oral NSAIDs and pneumonia in comparison to topical diclofenac (HR 3.49, 95% CI 1.59 – 7.66), while incorporating the first 5 days provided an effect estimate that was similar to the main analysis **(Figure 4)**. The hdPS-matched analysis results were similar to those of the main analysis (**Figure 4)**.

**Figure 4:**
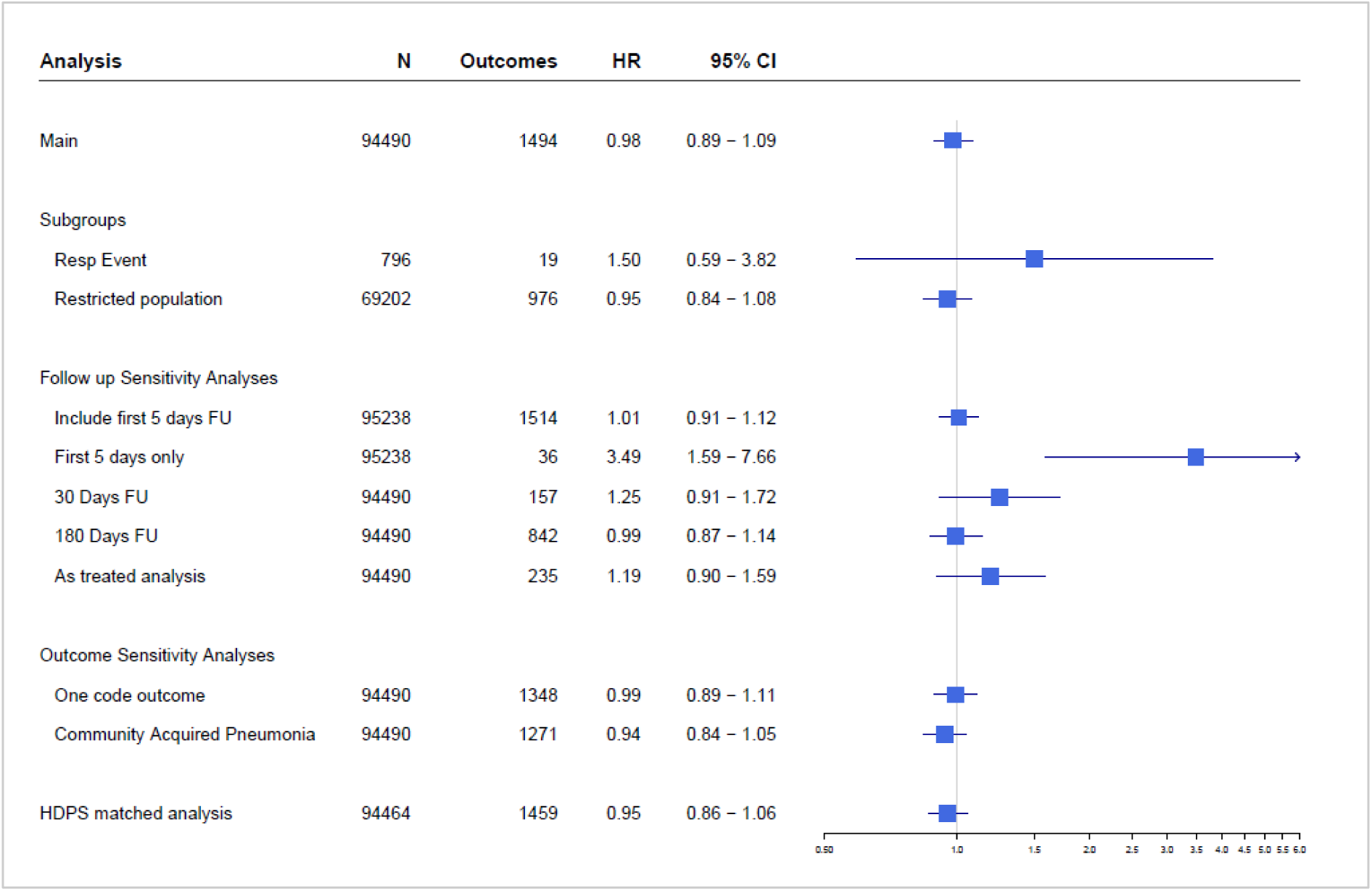
Forest plot demonstrating hazard ratios and 95% CI for the main NSAIDs-pneumonia analysis and all subgroup and sensitivity analyses. Main exposure was oral NSAIDs in osteoarthritis, comparator was topical diclofenac in osteoarthritis.

## Additional Analyses

In a general population, we found that new-users of non-selective NSAIDs had a 13% increased hazard of gastro-intestinal bleeding after propensity score matching relative to new-users of celecoxib, although the 95% confidence intervals included the null (HR 1.13, 95% CI 0.99 – 1.28). In a population restricted to those aged ≥60 years the risk increased to 27% (95% CI 1.09 – 1.49) **(See APPENDIX 3 for full results)**.

## Discussion

We conducted two separate cohort studies to further evaluate signals that we generated in a large-scale hypothesis-free screening study.^3^ In the heart failure example, we did not find strong evidence to support an increased risk of heart failure (14%, 95% CI 0.90 – 1.43) in 506,182 patients treated with clarithromycin or azithromycin when compared to 506,182 similar patients treated with amoxicillin/clavulanate. On the absolute scale, the difference in risk between treatment groups was small; translating to 4 additional cases per 100,000 person years in the macrolides group relative to the amoxicillin/clavulanate group.

In March 2018, the U.S Food and Drugs Administration published a drug safety communication advising caution when prescribing clarithromycin in patients with coronary heart disease because of growing evidence to support an increased risk of cardiovascular or all-cause mortality many years later.^25^ Long term data have come from CLARICOR, a randomized controlled trial from which 10-year follow up data demonstrate increased all-cause mortality in those randomized to clarithromycin *versus* placebo (HR 1.10, 95% 1.00 – 1.21).^6,7,22^ However, there is no biological explanation for this relationship. Furthermore, multiple observational studies in different health care settings and different clinical populations have produced conflicting results; some agreeing with CLARICOR^5,26,27^ and others not.^8,9^

Schembri *et al*. investigated cardiovascular risk associated with clarithromycin in two separate clinical populations; those with community acquired pneumonia and those with exacerbations of COPD. Their unadjusted results indicated no increased risk of heart failure in the pneumonia cohort but double the risk in the COPD cohort. The non-exposed group in the COPD cohort did not necessarily receive an antibiotic, thus potentially contributing to bias. A post-hoc analysis from a randomized trial of macrolides and fluoroquinolones *versus* beta-lactam antibiotics for the treatment of pneumonia found an association between erythromycin and heart failure (HR 1.89, 95% CI 1.22 - 2.91).^28^ The difference between this finding and our result may rest on differing study designs, exposures and included populations.

We found that the hazard ratio for heart failure was strongest in the first seven days of follow up, and weakest (crossing the null) when these seven days were removed from follow up. This possibly indicates a bias as truly incident heart failure may take a long time to manifest and diagnose; although we are uncertain of what the mechanism of bias might be. We are unconvinced of detection bias given that we achieved good balance on covariates even in the unmatched cohort. We included both myocardial infarction and arrhythmia as secondary outcomes because macrolides may be associated with these adverse events. ^5,29 30 31^ Heart failure may occur downstream of these outcomes. However, we found no evidence to support a relationship between either cardiovascular outcome and clarithromycin or azithromycin in our cohort. Thus, we cannot offer a plausible biological argument for the association between macrolides and heart failure, giving further weight to a conclusion of bias.

We did not find any evidence to support an association between oral NSAIDs and pneumonia (HR 0.98, 95% CI 0.89 – 1.09). The risk of pneumonia was strongest in the 5 days after exposure to oral NSAIDs, but weakened when these 5 days of follow up were removed from the analysis (main analysis) – suggesting protopathic bias. ^10-12^ Prior research has found that individuals treated with NSAIDs in the days preceding admission to hospital for pneumonia had more severe presentations, possibly due to altered immune responses and longer times to antibiotic treatment.^11,12^ Severe presentations increase the likelihood of pneumonia being diagnosed or detected.

Our studies have several strengths. In addition to our two substantive investigations, we also addressed a signal that is well known; gastrointestinal bleeding associated with exposure to NSAIDs, to demonstrate that our database and methods can faithfully reproduce signals we know to be true (**APPENDIX 3)**. We used a different database in a different country from the one that generated the signals in the original SSA. We used a new-user, active-comparator design with a comprehensive list of covariates in each analysis and propensity score matching to address measured confounding. Each analysis included large sample sizes. We carried out multiple sensitivity analyses to test our assumptions and definitions.

Our study also has limitations. We used a broad range of ICD-9 codes to define pneumonia as an outcome; however these codes have been found to have 71% positive predictive value when compared to chest x-ray.^15^ Nonetheless, our results were robust to alternative outcome definitions. Some residual confounding may have occurred in the heart failure example if for example an unmeasured covariate such as smoking was imbalanced between the groups, although our active comparator design with therapeutic alternative of amoxicillin/clavulanate achieved excellent balance on measured covariates and should help mitigate potential imbalances for unmeasured factors.^32^ We also used hdPS, which did not materially alter the results. Our data were derived from a commercial insurance claims database, which includes employer-based health insurance plans, thus our sample is not reflective of all populations. The association between macrolides and heart failure should be examined in a database with a population of higher mean age.

## Conclusion

We carried out an in-depth study of drug safety signals generated from a large-scale hypothesis-free SSA. We did not find strong evidence to support an increased risk of heart failure after exposure to macrolides and our range of sensitivity analyses suggest that some bias may be present, although we are uncertain of the mechanism. Further research is warranted to untangle this bias. Our findings for NSAIDs and pneumonia agree with previous findings from the literature and suggest detection or protopathic bias as an explanation for the original signal. Our study involving two independent investigations demonstrates how signals from large-scale screening activities can be efficiently examined using secondary claims data and how different databases and nuanced methods are paramount to refining safety signals.

## Data Availability

The raw data cannot be shared, however all diagnosis and drug code lists are readily available.

## Declarations

### Availability of data and material

*The raw data cannot be shared, however all diagnosis and drug code lists are readily available.*

### Code availability

*These analyses were conducted in the Aetion Evidence Platform^®^. Thus, code is not available but all decisions concerning cohort construction, advanced analysis plans, and generation of results are available in a transparent, regulatory grade paper trail.*

